# Wearable heart rate variability monitoring identifies autonomic dysfunction and thresholds for post-exertional malaise in Long COVID

**DOI:** 10.1101/2025.03.18.25320115

**Authors:** Twan Ruijgt, Anouk Slaghekke, Anneke Ellens, Kasper W. Janssen, Rob C.I. Wüst

## Abstract

**Objectives:** Patients with Long COVID experience disabling fatigue, autonomic dysfunction, reduced exercise capacity, and post-exertional malaise (PEM). Heart rate variability (HRV) can evaluate autonomic function and monitor overexertion, potentially helping to mitigate PEM. This study aimed to use continuous multi-day HRV recordings to monitor overexertion and study autonomic function in Long COVID.

**Method:** Heart rate and HRV were continuously measured in 127 patients with long COVID (43±11 years, 32% male) and 21 healthy controls (42±13 years, 48% male), and daily life activities tracked in a logbook. Participants underwent a (sub)maximal cardiopulmonary exercise test to determine heart rate at the first ventilatory threshold (VT1) to study HRV responses to exercise at different intensities.

**Results:** HRV was lower in patients with long COVID compared to healthy controls during various daily activities and sleep (p<0.027). HRV remained lower for 24 hours after exercise below, at or above VT1 in patients, but not in healthy controls (p=0.010). Nighttime HRV decreased with intense exercise and longer durations in patients with long COVID (p=0.018), indicative of exercise-induced diurnal disturbances of the autonomic nervous system in long COVID.

**Conclusion:** Heart rate variability, assessed by wearables, confirms autonomic dysfunction in patients with long COVID. The delayed recovery of the sympathovagal balance after exercise close and above to VT1 suggests that VT1 can be practically interpreted as a PEM threshold.

**Application:** These results confirm the applicability of wearables to assess autonomic function and manage overexertion in long COVID patients.

**Summary Box:** *What is already known on this topic:* Patients with long COVID often experience fatigue, autonomic dysfunction, and post-exertional malaise (PEM). HRV can be used as a non-invasive tool to measure autonomic function and recovery. Anecdotal evidence suggests lower HRV in patients with long COVID, but measurements are usually very short.

*What this study adds:* This study demonstrates that continuous HRV monitoring through wearables can effectively identify overexertion and autonomic dysfunction during daily activities in patients with long COVID. Patients with long COVID have a lower heart rate variability during sleep and HRV remained significantly lower for a longer period after moderate-to-heavy exercise, that is generally associated with the induction of post-exertional malaise.

*How this study might affect research, practice, or policy:* This study supports the use of wearables for assessing autonomic function and overexertion in daily life, helping patients with long COVID in pacing daily activities to mitigate symptoms of post-exertional malaise. HRV tracking after exercise shows that VT1 is a potential threshold for PEM. Sports physicians and physiotherapists can incorporate HRV biofeedback measures into pacing advice to patients. Additional research is needed to further investigate the effect of such an intervention.

## INTRODUCTION

Long COVID is associated with several symptoms, including severe fatigue, a lower exercise capacity, unrefreshing sleep, autonomic dysfunction, for example postural orthostatic tachycardia syndrome (POTS), and post-exertional malaise (PEM), which is the worsening of symptoms after physical or mental exertion above an unknown patient-dependent threshold [1–4]. Many patients experience significant setbacks following physical or mental exertion during daily life activities [1,5]. Autonomic dysfunction is implicated in the pathophysiology, although the exact cause remains unknown [6–8]. Suggested interventions for patients with long COVID involve heart rate pacing, keeping exercise intensity below this unknown PEM threshold [6,9]. Since this threshold is patient-dependent, a personalised approach in prescribing exercise and monitoring overexertion and recovery during daily activities is needed to improve patient well-being [5].

Heart rate variability (HRV) provides a non-invasive measure of autonomic function and can be used to monitor overexertion and recovery [8,10–12]. A commonly used method to determine HRV is expressing HRV as the root mean square of successive differences between RR-intervals (rMSSD) [13,14]. The HRV rMSSD is a marker for the sympathovagal balance and particularly the vagal activity of the parasympathetic nervous system [12,13]. Typically, higher HRV indicates better overall health, although significant inter-individual variability exists [15], partly related to sex and age-related reductions [16,17]. Given that a lower HRV is associated with diminished parasympathetic activity and increased stress, it may be a valuable marker for assessing autonomic function and recovery [18].

Patients with long COVID have lower HRV values at rest in the time-domain [8,19] or during sleep in the frequency domain [8], suggestive of parasympathetic inhibition and autonomic dysfunction. However, some studies even report higher HRV in the time-domain in long COVID patients [20,21]. A recent literature overview confirmed a lower HRV in patients with long COVID, although methodological quality was poor and measurements were very short and not related to specific daily life activities or sleep [22]. These findings suggest autonomic dysfunction, characterised by diminished parasympathetic activity, which may be a key feature in long COVID patients [8,19], but how HRV is related to the pathophysiology of ppost-exertional malaise in long COVID is currently unknown. Indeed, HRV in patients with myalgic encephalomyelitis/chronic fatigue syndrome (ME/CFS) remained lower shortly after exercise, while HRV increased in healthy controls [23].

Exercise intensity and duration significantly modulate the responses of the (para)sympathetic autonomic nervous system during exercise [24]. The parasympathetic nervous system is typically more active at rest and during low-intensity exercise, while sympathetic activity increases during high-intensity or prolonged exercise near or above VT1. How heart rate variability responds to acute exercise, and what the recovery responses after exercise are in patients with long COVID is, however, currently unknown.

Gaining a better insight into HRV responses during sleep, daily life activities and the recovery after moderate-to-heavy exercise will not only provide more fundamental insights into the pathophysiology of PEM and long COVID, but also help patients with structuring daily activities and prevent overexertion and PEM. Multiple 24-hour recordings during sleep and daily life activities, encompassing the recovery after periods of moderate-to-heavy exercise will provide a better understanding of the diurnal adjustments in HRV in long COVID patients. The aim of this study was therefore to measure HRV during sleep and daily life activities over multiple days in patients with long COVID to study HRV during specific daily life activities, with particular focus on the recovery responses of HRV after moderate and heavy exercise above VT1 in healthy controls and patients with long COVID.

## METHODS

An extended methods section is provided in the supplemental information.

### Participants

This retrospective study included patients with long COVID who were referred to a sports and exercise medicine clinic (DeSportarts, Utrecht, the Netherlands). Inclusion criteria were 1) minimal one SaRS-CoV-2 infection, as diagnosed by a general practitioner, and 2) being treated by a general practitioner for long COVID-related symptoms, including severe fatigue, brain fog, sleep disturbances, and particularly post-exertional malaise. A control group consisted of participants matched for age and sex to the long COVID group. Control participants could have had a SaRS-CoV-2 infection, but without long COVID symptoms. Exclusion criteria for the control group were chronic illnesses and related medication use or being an elite athlete. All patients gave Informed consent to use the collected data for this research. The medical ethical committee of the Amsterdam UMC waived ethical oversight according to local legislation.

### Determination of exercise intensity domains

There is currently no scientific consensus about the PEM threshold, but anecdotal, clinical evidence suggests that the first ventilatory threshold (VT1) might provide some indications of such a threshold [25]. VT1 reflects the point where the body begins to accumulate low levels of lactate in the blood.

Current optimal treatment to avoid PEM in patients with long COVID includes avoiding overexertion, and as such, long COVID participants were instructed to avoid exercise above the heart rate at the first ventilatory threshold. This was determined in a submaximal exercise test on a bike, three to six weeks prior to the HRV assessment. Furter details can be found in the supplemental file. VT1 was determined using the V-slope and ventilatory equivalent methods and was assessed by two experts. Heart rate at the first ventilatory threshold was defined as the patient-specific PEM threshold. Heart rate pacing was not advised in the control group.

Post-hoc analyses allowed us to classify long COVID patients based on daily-life-impairments into mild and moderate patients. We reasoned that ∼15 ml O_2_.kg^-1^.min^-1^ is needed for daily life activities [26]. Patients with VT1 values >15 ml O_2_.kg^-1^.min^-1^ were classified as mildly impaired, and patients with VT1 values <15 ml O_2_.kg^-1^.min^-1^ were classified as moderately impaired.

### Heart rate variability assessments and data analysis

All participants were instructed to wear a Bodyguard 3 device (Firstbeat, Finland), a clinical grade *ECG*-based HRV wearable, to measure heart rate variability (HRV). The device was attached using electrodes (CardinalHealth, Kendall, 57×34mm) and placed beneath the right collarbone and below the left chest (Figure 1A). The Bodyguard uses a 1 channel ECG at 256 Hz, and measures RR-intervals with 1-ms intervals. Participants wore the bodyguard continuously for 3-7 consecutive days, excluding water contact (shower, bath etc.). Collected data was uploaded anonymously into the Firstbeat cloud. Participants documented their daily life activities in the Firstbeat Life app. Retrospectively, activities were categorised in strict domains: sleep, eating, travel, relaxation, work, walking, exercise (light, moderate or intense), household chores, reading, relaxing exercises, getting ready, and watching TV. HRV rMSSD, averaged over a 5-minute period by the Firstbeat software, was used for the analysis of the daily life activities. HRV data for each category was averaged per participant across all measurement days to provide a personal, averaged HRV per daily life activity.

**Figure 1.**
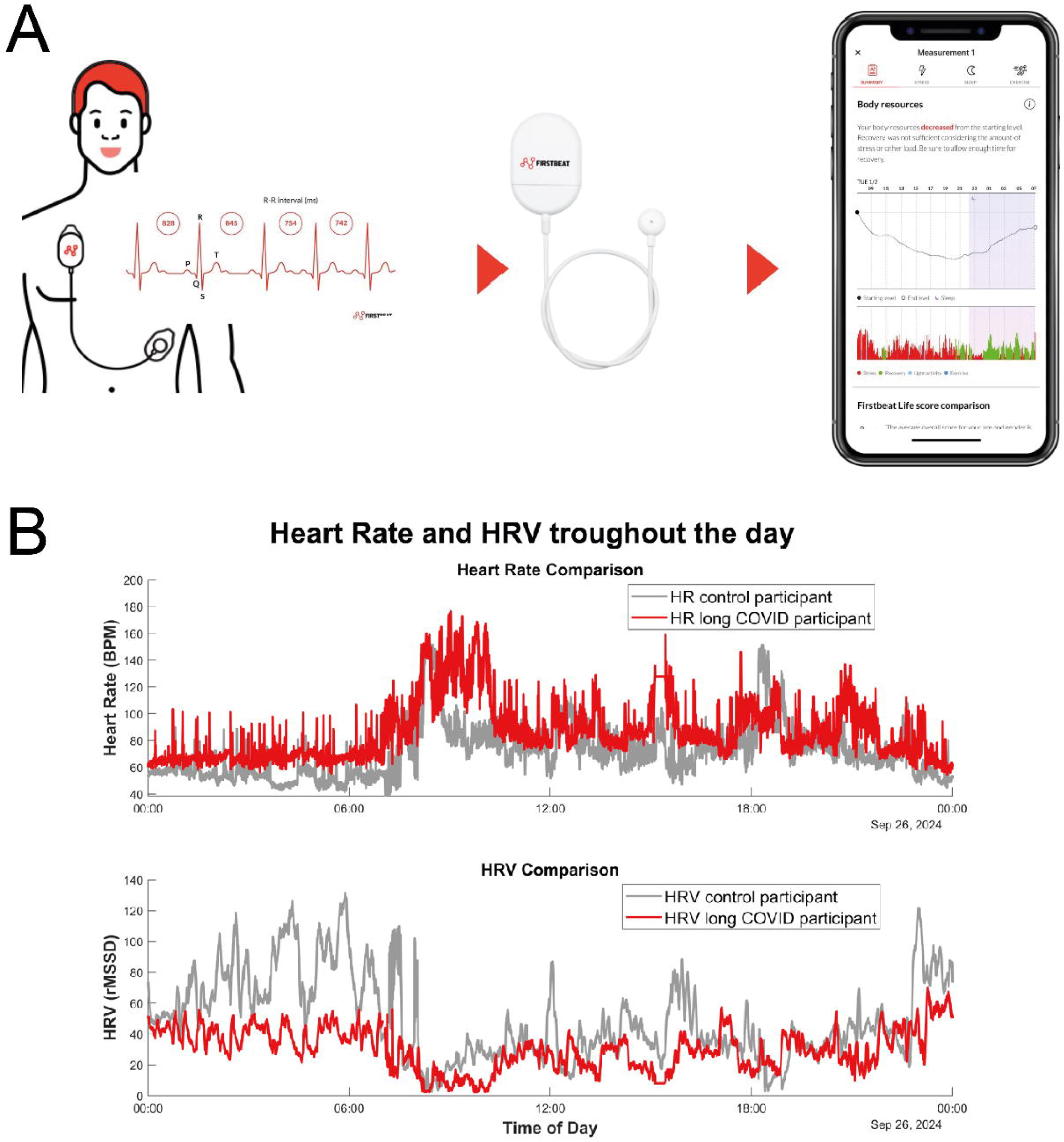
**A:** Schematic overview of the measurement. The Firstbeat Bodyguard 3 device is attached beneath the right collarbone and left chest area. RR-intervals are recorded and stored in a digital database, and results can be viewed in an app. Data can be extracted for research purposes. **B:** Typical example of heart rate and heart rate variability (HRV) recordings during one day for one healthy control and one patient with long COVID, with identical age, sex and BMI. HRV was calculated as the root mean square of successive differences between RR-intervals (rMSSD). When heart rate increased, HRV typically decreases. Healthy controls typically had more daytime fluctuations in HRV rMSSD compared to patients with long COVID.

For the HRV after exertion analysis, data analysis was conducted using Matlab R2023b. Details are specified in the supplemental file. A typical figure of HRV and HR throughout the day in both groups is shown in Figure 1B.

#### Heart rate variability during recovery after exercise

Exercise intensities were categorised into three levels: mild (80-90% of HR at VT1), moderate (90-100% of HR at VT1), and intense (HR above value at VT1). For every hour after exercise completion (up to 24 hrs), a 15-minute average HRV was computed. Exercise was defined as maintaining a heart rate above the predefined exercise intensities for at least 20 minutes. Because not all healthy controls underwent a submaximal exercise test, heart rate at VT1 was predicted using 75% of the age-predicted maximum heart rate in a subset of the healthy controls.

Additionally, the percentile difference between the highest nighttime HRV across all measurement days, and the HRV during a single night was calculated per participant to evaluate the impact of exercise-induced stress on nighttime HRV during sleep. Therefore, the total time spent in each exercise category during the day was classified as short (<20 min), medium (20-60 min) and long (>60 min).

### Statistical analysis

Statistical analysis was performed using R (version 4.4.1). Descriptive data were reported as mean ± standard deviation, unless noted otherwise. Normality was checked using Shapiro-Wilk tests and visually inspected with Q-Q plots. Significance was accepted at p<0.05. Post-hoc comparison was performed using Bonferroni.

Differences in HRV rMSSD during daily life were analysed using a non-parametric rank-based regression model (R-fit), with group (control vs. long COVID) as the independent factor and age and sex as covariates, given their established correlations with HRV (Supplemental Figure 1). The normally distributed heart rate data were analysed using a linear model with age and sex again as covariates. Differences in recovery HRV rMSSD after exercise were analysed using a rank-transformed mixed linear model to account for repeated measures, with severity group (moderately, mildly impaired, control), timepoints (recovery hours), and exercise intensity (80-90%, 90-100%, >100% of heart rate at VT1) as fixed effects, and age and sex as covariates. For the exercise-induced alterations in nighttime HRV, we analysed the mean relative change in nighttime HRV compared to the best night using a linear mixed-effects model. Group (long COVID vs control) and time spent per exercise intensity during the day (<20 min vs >60 min) were included as fixed effects. If exercise was performed on multiple days, the mean HRV of each participant was calculated.

## RESULTS

This study included 121 long COVID patients and 21 control individuals (Patient characteristics in Table 1). Long COVID participants had on average a higher BMI compared to the age- and sex healthy controls. Patients with long COVID had a lower power output and oxygen uptake consumption (*V̇* O_2_) at VT1 compared to controls (p<0.001). Long COVID patients were classified as moderately impaired, with VT1 <15 ml.kg^-1^.min^-1^ (which is the oxygen consumption needed for daily life activities; N=34, 24% male, 43±11 years, *V̇* O_2_: 13.1±1.2 ml.kg^-1^.min^-1^) or mildly impaired (VT1 >15 ml.kg^-1^.min^-1^; N=87, 36% male, 41±10 years, *V̇* O_2_: 21.6±5.1 ml.kg^-1^.min^-1^).

**Table 1.** Participant characteristics. Data is shown as mean±SD.

|  | **Healthy**<br>**Control** | **long COVID** | ***p-value*** |
| --- | --- | --- | --- |
| Number of participants | 21 | 121 |  |
| *Daily life* |  |  |  |
| Age (years) | 42 ± 13 | 42 ± 11 | 0.87 |
| Sex (male %) | 10 (47.6%) | 35 (31.0%) | 0.17 |
| Height (cm) | 178 ± 9 | 174 ± 10 | 0.14 |
| Weight (kg) | 73.4 ± 13.0 | 77.6 ± 14.5 | 0.22 |
| BMI (kg.m<sup>-2</sup>) | 23.1 ± 3 | 25.7 ± 4.7 | 0.02 |
| *Submaximal exercise test* |  |  |  |
| Number of participants | 9 | 121 |  |
| Age (years) | 41 ± 15 | 42 ± 11 | 0.74 |
| Sex (male) | 4 (44.4%) | 35 (31.0%) | 0.46 |
| BMI (kg.m<sup>-2</sup>) | 23.9 ± 2.6 | 25.7 ± 4.7 | 0.31 |
| Power output at VT1 (W) | 183 ± 59 | 100 ± 44 | <0.001 |
| $\dot{V}O_2$ at VT1 (ml.kg<sup>-1</sup>.min<sup>-1</sup>) | 28.6 ± 6.1 | 19.2 ± 5.8 | <0.001 |
| RER at VT1 | 0.95 ± 0.04 | 0.95 ± 0.04 | 0.74 |

### Adverse effects from the CPET

Three patients from this cohort experienced PEM for more than 5 days after the submaximal CPET. More than 5 days was considered an adverse event, because of the prolonged negative impact on daily life.

### HRV during daily life activities

HRV rMSSD was lower in the long COVID group during sleep, eating, travelling and the uncategorised (and combined) activities (*p=0.005-0.027*; Figure 2, Supplemental Table 1 and 2). Notably, 45% of reported activities were classified under this non-specific category, as these activities consisted of those that did not fall directly into one of the 12 predefined categories (e.g. shopping) or were a combination of two activities (e.g. eating in front of TV). A general negative correlation between HR and HRV is shown in Supplemental Figure 2. Heart rate was significantly higher during sleep and the uncategorized activities in patient with long COVID, but no such difference was observed during eating or traveling. The daily life activities with higher absolute heart rates and subsequently lower HRV values, such as exercise, showed no significant differences in HRV, likely due to a more pronounced sympathetic nervous system activity.

**Figure 2.**
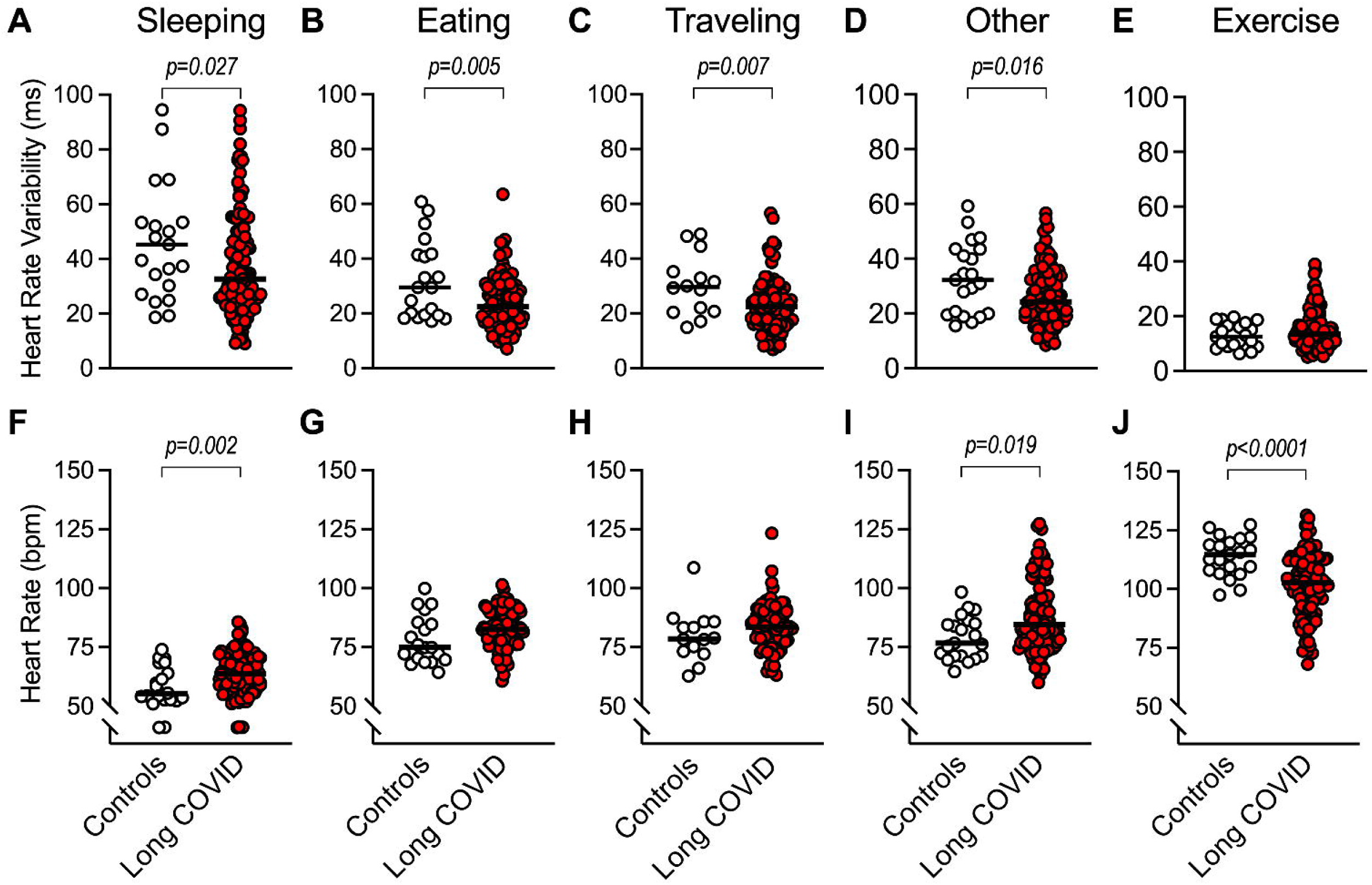
Heart Rate (Variability) during daily life activities. HRV (**A-E**, measured as rMSSD) and heart rate (**F-J**, in bpm) are shown during different daily life activities: sleeping (**A, F**), eating (**B, G**), traveling (**C, H**), uncategorized activities (**D, I**), and exercise (**E, J**). HRV was significantly lower in patients with long COVID compared to healthy controls during sleep (p=0.027), eating (p=0.005), traveling (p=0.007), and uncategorized activities (p=0.016). No difference in HRV was observed during exercise (**E**), despite significantly higher heart rates in controls (p<0.0001, **J**). In long COVID patients, heart rate was significantly higher during sleep (p=0.002, **F**) and uncategorized activities (p=0.019, **I**), while no significant differences were observed during eating (**G**) or traveling (**H**). Median values (horizontal bars) and individual data points are displayed.

### Recovery of HRV after exercise cessation

As pacing advice is the current optimal treatment for patients to avoid PEM, we first studied how many patients recorded heart rates above the VT1. In 43% of patients with long COVID we recorded heart rates above the anaerobic threshold during daily-life activities. Thus, the adherence to the heart rate pacing advice given after the CPET was 57%. We stratified exercise into three exercise categories: mild (heart rate at 80-90% VT1), moderate (90-100% VT1), or intense (>100% VT1) exercise, and we studied the recovery of the heart rate variability after exercise cessation. For this analysis, we made a subdivision between mildly and moderately impaired patients based on oxygen consumption at VT1. Figure 3 illustrates the time course of HRV during the recovery period after mild, moderate, and intense exercise. Across all exercise intensities, patients with long COVID had significantly lower HRV values compared to healthy controls (p=0.010), with no differences between mildly and moderately impaired patients. HRV reduced the most after intense exercise in the mildly affected patients (p<0.0001), but not in the moderately impaired patients where HRV was consistently low. HRV increased relatively rapidly after exercise cessation in healthy controls (3 to 6 hours post-exercise; Figure 3 blue markers). This delayed recovery-associated increase in HRV (which is independent of heart rate) was significantly faster in healthy controls compared to mildly and moderately impaired long COVID patients (p=0.002). While HRV increased relatively rapidly after mild intensity exercise in mildly affected patients (5-6 hours), the HRV recovery was delayed after exercise above VT1 (until 9 hours) in the mildly affected patients. HRV did not increase, relative to the exercise itself, until 10-13 hours in the moderately impaired patients at various exercise intensities above 80% of VT1.

**Figure 3.**
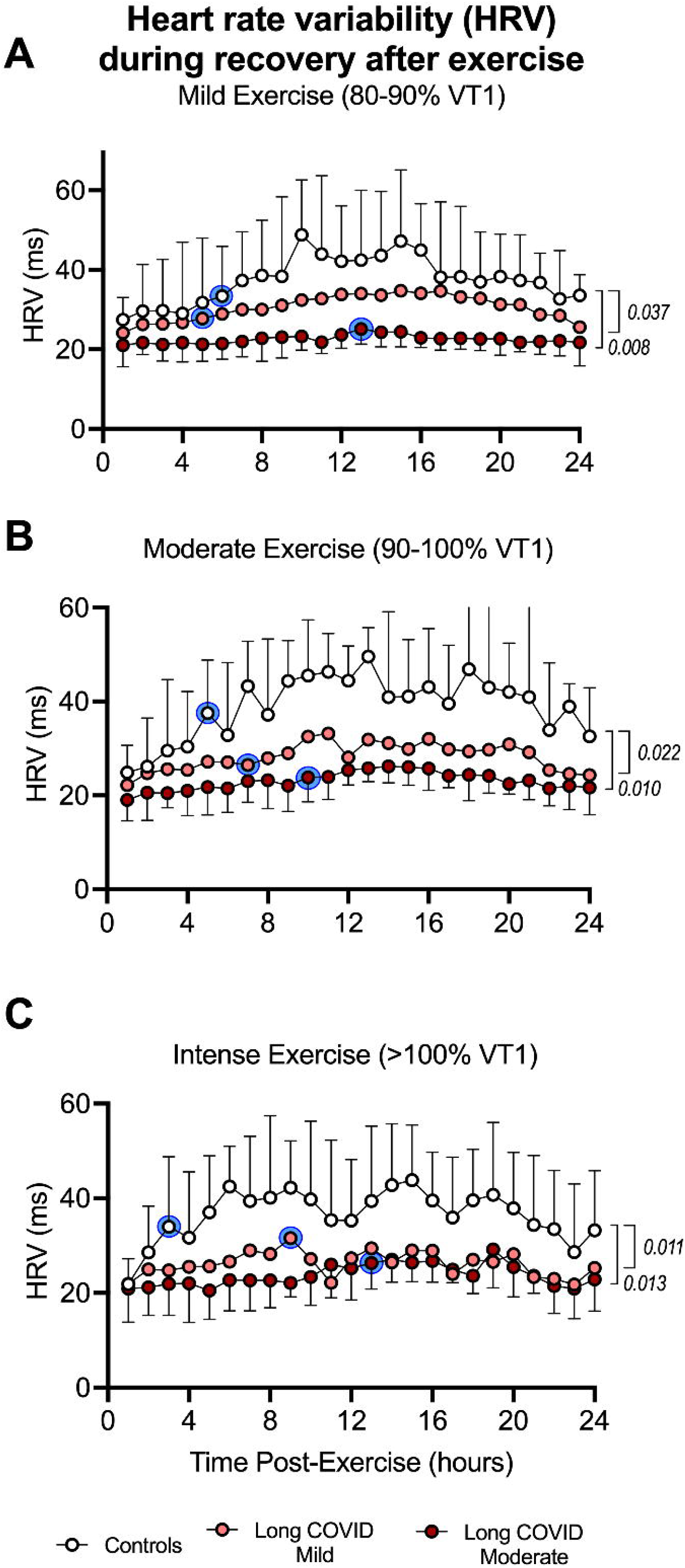
Recovery of heart rate variability (HRV) after exercise cessation. The time course of heart rate variability (HRV, as measured as rMSSD) for 24 hours after cessation of exercise at mild (**A:** 80-90% VT1), moderate (**B:** 90-100% VT1), and intense (**C:** >100% VT1) intensity. Healthy controls (white) had higher HRV compared to patients with mild (pink) and moderate (dark red) long COVID. Blue-shaded circles indicate the time points at which HRV values became significantly higher (p<0.05) compared to the 1-hour post-exercise baseline within each group. Data points represent the median HRV values for each group at each time point, with error bars indicating the 95% confidence intervals.

### Nighttime HRV following exercise

We reasoned that the delayed recovery time course in HRV in patients would eventually affect nighttime HRV. Indeed, nighttime HRV was consistently reduced following exercise, with differences across exercise durations and intensities (Figure 4A). The nighttime HRV of healthy controls and patients reacted differently to the day-time exercise bout. Nighttime HRV in the controls was higher, and did not significantly depend on exercise duration or intensity. In patients however, increasing exercise duration and intensity both resulted in significantly lower HRV values during the subsequent night. The interaction between exercise duration and intensity (p=0.010) suggests that the extent of HRV reduction during the subsequent night depends on the combination of exercise duration and intensity (Figure 4B). When considering changes in nighttime HRV relative to an individual’s best night, both exercise intensity and duration significantly influenced nighttime HRV reductions in the long COVID group. Specifically, engaging in more than 60 minutes of exercise, regardless of intensity, led to a greater reduction in nighttime HRV compared to performing less than 20 minutes of exercise at the same intensity (p<0.033; Figure 4B). Additionally, a significant difference was observed between mild and intense exercise when performed for less than 60 minutes, with intense exercise causing a greater reduction in nighttime HRV (p<0.038). The significant interaction effect between time and intensity (p=0.018) further underscores the combined impact of both factors on nighttime HRV recovery, particularly in patients with long COVID. Together, these results illustrate that exercise with increasing intensity and/or duration causes a delayed adjustment of HRV in patients with long COVID, likely related to disturbances in parasympathetic response in patients with long COVID. As even short duration exercise at intensities above VT1 were associated with abnormal diurnal adjustments of HRV in patients.

**Figure 4.**
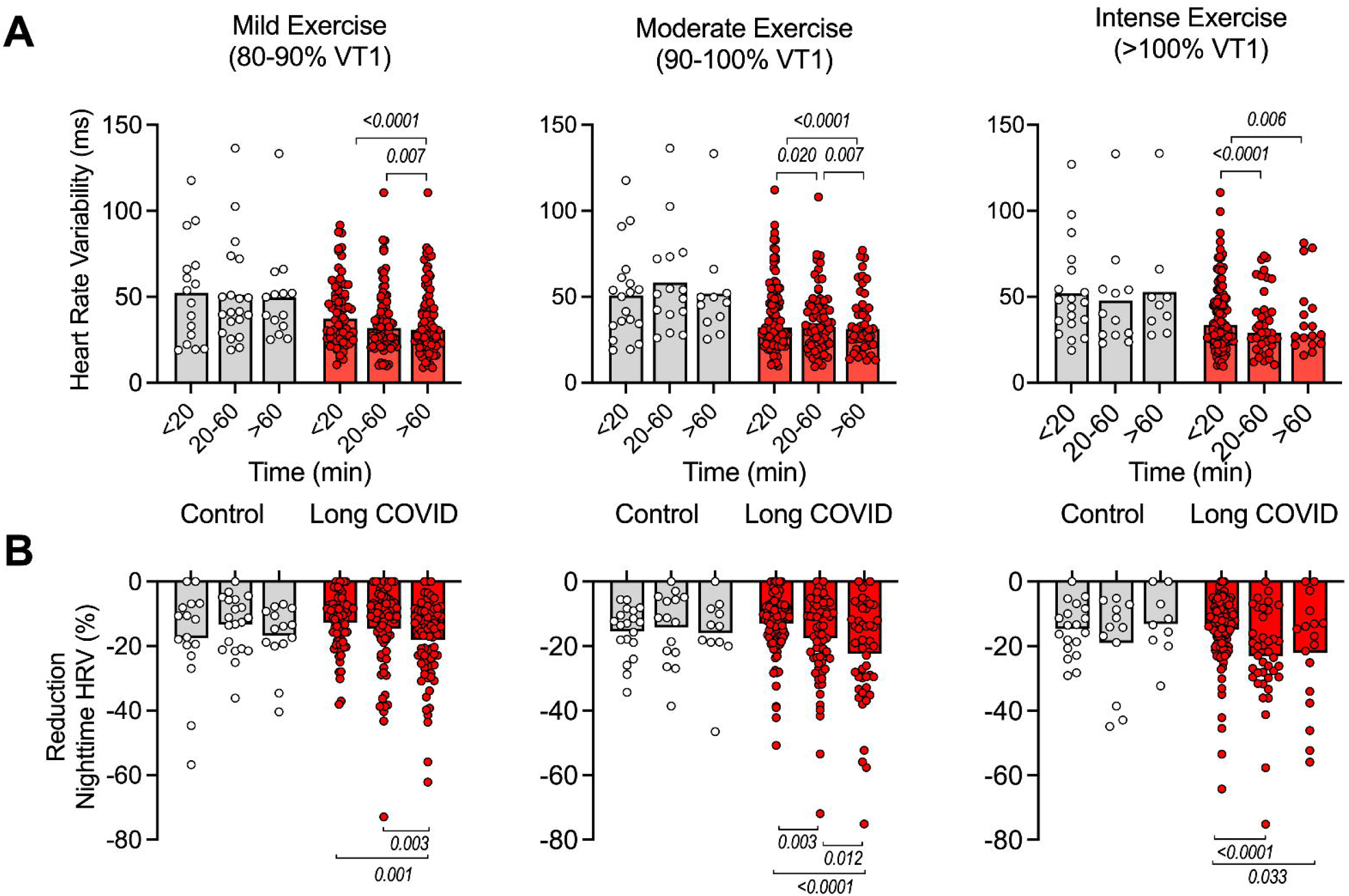
Heart rate variability during post-exercise night. **A:** Heart rate variability (HRV) during the night following mild (80-90% of VT1), moderate (90-100% of VT1), and intense (>100% of VT1) daytime exercise. HRV was lower, only in patients, following exercise with increasing duration (around the VT1) and exercise intensity. **B:** The relative change (%) in nighttime HRV (measured as rMSSD) is shown following mild (80–90% of VT1), moderate (90–100% of VT1), and intense (>100% of VT1) daytime exercise. Also the relative reduction was lower in patients, following exercise with increasing duration and exercise intensity. Exercise durations were divided into three categories: <20, 20-60 and >60 min. Bars represent group means, and individual data points are shown.

## DISCUSSION

The aim of this real-life study was to use HRV during daily life activities to study autonomic function and the recovery responses to exercise in patients with long COVID. The adherence to the heart rate pacing advice was substantial, 57% of patients did not exceed their VT1 threshold during the HRV measurement. We observed that HRV was lower in long COVID patients in multiple daily life activities compared to healthy controls. HRV in patients with long COVID remained low during the recovery period after mild, moderate, or intense exercise when compared to healthy controls. Lastly, long-lasting exercise around or above the VT1 resulted in a lower HRV during the subsequent night in patients with long COVID.

### Autonomic dysfunction in long COVID

Our findings of a lower HRV in patients with long COVID are consistent with earlier research [16–18]. The lower HRV during sleep and many daily life activities in patients compared to healthy controls suggest that long COVID patients may experience significant autonomic dysfunction, characterised by reduced parasympathetic activity and/or an increased sympathetic activity throughout a large proportion of their daily life, and during the night. The lower HRV observed during sleep, a critical period for restoration, can limit the ability of the body to recover from daily stressors. These alterations can be linked to the unrefreshing sleep that patients with long COVID experience.

The pathophysiology of autonomic nervous system dysfunction in long COVID is unclear. The autonomic modulation of the (para)sympathetic activity during exercise is partly mediated by the release of catecholamines. Autoantibodies against various catecholamine receptors involved in the autonomous nervous system have been observed in patients with long COVID and these correlate with symptom severity [27]. Some new studies have indicated a direct consequence of the viral infection for brain stem function [28]. Whether autonomic nervous system dysfunction is a response to other neuropathophysiological alterations [29] or autoimmunity [30] needs further study. Also, how acute exercise modulates the response of the immune system to the viral persistence or autoantigen presentation, and how this links to the onset of post-exertional malaise in patients with long COVID is currently unknown.

### Recovery after moderate to heavy exercise

As the parasympathetic nervous system is more active during lower-intensity exercise, the first ventilatory threshold demarcate a threshold for sympathetic nervous system activation as measured using HRV recovery [31]. Recovery of parasympathetic activity is typically delayed (up to 60-120 minutes) after exercise above VT1, even in highly-trained athletes [31]. In our real-life study, we observed that in healthy individuals, HRV values recover within a few hours after the completion of exercise, as the ratio of sympathetic and parasympathetic activities shift back towards mainly parasympathetic activities, which aids in recovery. However, this increase in HRV during the recovery of exercise was delayed for a significantly longer time during in long COVID patients and did not rise back to the same values as in healthy controls, particularly following exercise at intensities above the VT1, particularly in the moderately impaired patients. This blunted HRV recovery after exercise suggests an insufficient or delayed response in the reduction in sympathetic nervous system activity, and/or an increase in the parasympathetic activity after exercise. These inabilities to return heart rate variability back to baseline values indicate that exercise recovery is impaired in long COVID, and more negatively affected in moderately impaired patients. This could contribute to a prolonged physiological stress and a delayed recovery period. Above the VT1, the sympathetic activity and stress hormone concentrations (i.e. cortisol and catecholamine) typically increase [32], but we did not measure stress hormones concentrations and time courses during and after high-intensity exercise in patients with long COVID.

We even observed lower night-time HRV following day-time exercise with a long duration and intensity, suggesting that moderate-to-intense physical exercise continues to suppress parasympathetic activity during subsequent sleep resulting in a reduced recovery.

Our findings using real-life data from patients provide further insights into the pathophysiology of post-exertional malaise (PEM), in which physical, cognitive or mental exertion leads to worsening of symptoms and longer recovery times [1]. The acute initiating PEM-inducing response to exercise in patients is unknown. Since PEM symptoms are not limited to skeletal muscle-associated abnormalities [1], we provide evidence that acute exercise leaves an autonomous nervous system signature that could, at least in theory, initiate multi-organ symptomology including unrefreshing sleep and worsening of other symptoms. The underlying pathophysiology of PEM and the link with abnormal autonomous nervous system activities is unknown, as we do not have recordings of PEM episodes in this real-life study. As such, more controlled studies need to be performed to further understand this.

Currently, no curative treatment for PEM is available, and patients are advised to limit intense exercise above the PEM-threshold. This intensity of the PEM-threshold is currently unknown, but as HRV recovery responses were particularly disrupted with exercise of a longer duration (>60 min) and at intensities near VT1, we hypothesize that the PEM-threshold occurs near VT1. Anecdotal evidence from patients with long COVID suggests that this threshold is time- and patient-dependent, and is not only caused by muscular activities, but also by cognitive and mental exertion. As VT1 is mechanistically related to altered peripheral lactate production, this should be interpreted as a practical marker for the PEM-threshold. VT1 is also associated with increased sympathetic activity and release of cortisol and catecholamines (cortisol and (nor)adrenaline) into the blood stream [32], but it is unknown whether the increase in sympathetic activity or the circulating catecholamines, or other hormonal alterations provide a more mechanistic underpinning of the PEM-threshold. Whether an abnormal physiological stress response, or disturbed relaxation response underlies the acute onset of PEM deserves further study.

### Practical applications

#### Biomedical basis for long COVID

Our findings provide further evidence of physiological alterations during and after physical exercise in patients with long COVID that are distinct from healthy controls. Autonomic dysfunction and impaired recovery in HRV in long COVID patients contribute to our understanding of abnormal exercise responses in long COVID.

#### Application of wearables

Wearables, such as Firstbeat Bodyguard 3, effectively monitor heart rate variability (HRV) in patients and controls during sleep, daily life activities and recovery after exercise. Patients can use such devices to aid in identifying stressful moments, overexertion and to recognise personalised patterns that influence nighttime recovery. Adjusting daily life activities appropriately can help in mitigating symptoms and improving overall well-being. As long COVID symptoms, recovery after exercise, and resting HRV are individual, such pacing strategies clearly require a personalised approach.

#### Guidelines for exercise intensities

The impaired HRV recovery seen in long COVID patients emphasises the importance of managing exercise intensities and duration carefully, as overexertion not only impacts immediate recovery, but can also disrupt sleep and subsequent recovery. Our results indicate that physical exercise with increasing intensity and duration significantly reduces subsequent HRV during the night, impairing sleep recovery. Sports physicians can conduct a submaximal exercise test to determine the heart rate at VT1, which helps patients in providing a usable marker for the PEM-threshold during physical exercise. Continuous heart rate and HRV monitoring throughout the day can be done using wearables, which help improve recovery and mitigate symptoms of post-exertional malaise.

### Limitations

This real-life study using HRV monitors in patients has several limitations. Firstly, the real-life setting of this study meant that a strict experimental protocol could not be applied. A wide range of (combined) daily life activities were recorded in the logbook, which increased subject-to-subject variability. Some activities were reported by only a small number of participants, limiting the statistical power. Current best treatment for patients with long COVID include the avoidance of very intense PEM-inducing exercise, and as such, we do not have controlled PEM induction in this study. The goal of heart rate pacing below VT1 was to prevent PEM. Without heart rate pacing, the exertion in the long COVID group might have been higher leading to even more pronounced differences compared to healthy controls. Symptom questionnaires would have allowed us to correlate abnormal HRV recovery after exercise with PEM symptomology. The severity and time since the initial COVID-19 infection in long COVID participants was unknown, which may have influenced HRV values. We acknowledge the relatively small control group.

### Future research

Controlled trials with PEM induction and continuous follow-up of heart rate, heart rate variability and other parameters related to the autonomous nervous system would provide a more mechanistic insight into the underlying pathophysiological mechanisms underlying autonomous and endocrine dysregulation during exercise in patients with long COVID, and the putative link with the threshold for post-exertional malaise. A second line of research relates to the practicability of HRV for pacing. Clinical trials on the (lack of) effectiveness of exercise training, pacing and use of wearables in patients with long COVID are urgently required.

## CONCLUSION

In conclusion, here, we provide evidence that heart rate variability is lower during daily life activities in patients with long COVID and remains abnormal after exercise. The measurement of HRV is an easy-to-use non-invasive biomarker of exertion, that can be used to help with limiting overexertion and preventing subsequent induction of PEM in patients with long COVID, and likely also patients with post-acute infectious syndrome (PAIS) or myalgic encephalomyelitis / chronic fatigue syndrome (ME/CFS) who also suffer from PEM. HRV can be easily measured using wearables during daily life activities in patients, and can also be used for future clinical intervention therapies on treating or reducing PEM in patients.

## Data Availability

All data produced in the present study are available upon reasonable request to the authors

## Contributors

TR, RCIW and KWJ designed the study, all were involved in the data collection, analysis and interpretation; TR drafted the manuscript; all were involved in revision of the final version.

## Funding

This study was funded internally by DeSportarts and Vrije Universiteit Amsterdam with support from the ZonMw “Consortia voor biomedisch cohortonderzoek naar ME/CVS 2022” and “Biomedische kennishiaten post-COVID 2024”.

## Acknowledgements

The authors want to thank patient representatives for insightful discussions.

## Competing interests

None declared.

## Patient and public involvement

Patients with Long COVID were consulted in the design, and data interpretation of this project.

## Notes

### Competing Interest Statement

The authors have declared no competing interest.

### Author Declarations

The medical ethical committee of the Amsterdam UMC waived ethical oversight according to local legislation.

