## Supplementary material for "Wearable heart rate variability monitoring identifies autonomic dysfunction and thresholds for post-exertional malaise in Long COVID": Supplemtal file

**Extended methods**

**Participants**

This retrospective study included patients with long COVID who were referred to a sports and exercise medicine clinic (DeSportarts, Utrecht, the Netherlands). Inclusion criteria were 1) minimal one SaRS-CoV-2 infection, as diagnosed by a general practitioner (including PCR testing), and 2) being treated by a general practitioner for long COVID-related symptoms, including severe fatigue, brain fog, sleep disturbances, and particularly post-exertional malaise. A control group consisted of participants matched for age and sex to the long COVID group. Control participants could have had a SaRS-CoV-2 infection, but without long COVID symptoms. Exclusion criteria for the control group were chronic illnesses and related medication use or being an elite athlete. All patients gave Informed consent to use the collected data for this research.

**Determination of exercise intensity domains**

There is currently no scientific consensus about the PEM threshold, but anecdotal, clinical evidence suggests that the first ventilatory threshold (VT1) might provide some indications of such a threshold. VT1 marks the limit between the slight and moderate intensity of exercise and exercising around this threshold allows stimulating aerobic metabolisms while above VT1 blood lactate and pH start to increase and decrease, respectively.

Current optimal treatment to avoid PEM in patients with long COVID include avoiding overexertion, and as such, long COVID participants were instructed to avoid exercise above the heart rate at the first ventilatory threshold. This was determined in a submaximal exercise test on a bike, three to six weeks prior to the HRV assessment. Termination criteria were pre-determined in consultation with the participant. If serious PEM symptoms were anticipated, the test was terminated after reaching the first ventilatory threshold (VT1), which was the case for most Long COVID patients. Otherwise, the test continued until the second ventilatory threshold (VT2) or maximal exhaustion. The exercise test began with 2 minutes of rest on the ergometer followed by a 4-minute baseline cycling period, followed by a ramped, linear increase in power output at a rate of 10-20 W.min^-1^ in patients. 30W/min for healthy women and 40W/min for healthy men. Participants were instructed to maintain their cadence above 60 revolutions/minute. Throughout the test, ECG, heart rate, blood pressure, O_2_ saturation levels and Borg exertion levels were monitored. Pulmonary gas exchange and ventilation were measured on a breath-by-breath basis (Cosmed Quark CPET; Cosmed, Rome, Italy), and displayed during the test. VT1 was determined using the V-slope and ventilatory equivalent methods ($\dot{V}$_E_/$\dot{V}$O_2_) and was assessed by two experts. Heart rate at the first ventilatory threshold was defined as the patient-specific PEM threshold. Heart rate pacing was not advised in the control group.

**Data analysis of heart rate variability after exercise**

For the HRV after exertion analysis, data analysis was conducted using Matlab R2023b. RR-intervals were filtered with a slightly adjusted version of HRVTool (v.1.07, <https://marcusvollmer.github.io/HRV/>). In short, a threshold-based beat correction algorithm was applied, using a 0.20s threshold corresponding to the 'medium-strong' filter of the Kubios software. RR-intervals deviating by more than 0.20s from the local average (median of 30 surrounding RR-intervals) were corrected using cubic spline interpolation. This threshold was adjusted based on mean heart rate over a 20-second period; i.e. a heart rate of 120 bpm resulted in a filter threshold of 0.1s. HRV was calculated using the root mean square of successive differences (rMSSD) from 300 RR-intervals. Heart rate was averaged over 20-second periods.

**Supplementary tables and figures**

**Supplementary Table 1.** Heart rate (HR) during daily life activities. Median [interquartile range] (number of participants).

|  | **Healthy controls** | **Long COVID** | *P* value |
| --- | --- | --- | --- |
|  | HR (beats.min^-1^) | HR (beats.min^-1^) |  |
| Sleep | 55.4 [52.7-64.0] (21) * | 63.8 [59.1-70.5] (119) * | *0.002* |
| House chores | 86.4 [79.4-95.9] (18) | 89.1 [81.8-95.4] (108) | 0.413 |
| Eating | 74.9 [70.2-84.9] (20) | 82.4 [77.9-90.0] (105) | 0.076 |
| Working | 76.1 [69.0-83.0] (20) | 78.8 (72.5-86.4) (82) | 0.930 |
| Traveling | 78.6 [73.7-85.2] (14) | 83.6 [78.0-9.2] (86) | 0.142 |
| Relaxing | 73.2 [62.4-79.1] (16) | 74.4 [68.4-83.9] (100) | 0.168 |
| Watching TV | 71.1 [64.6-76.4] (14) | 74.4 [69.1-79.9] (99) | 0.201 |
| Walking | 101.3 [82.5-103.6] (13) | 91.7 [83.8-100.1] (97) | 0.317 |
| Exercising | 114.6 [108.1-119.8] (21) * | 102.7 [92.6-111.9] (103)* | *<0.001* |
| Getting ready | 82.7 [70.3-89.8] (7) | 86.9 [80.5-93.8] (74) | 0.439 |
| Relaxing exercise | 58.4 [58.4-58.4] (1) | 78.9 [72.3-89.3] (58) | 0.273 |
| Reading | 73.9 [65.2-76.4] (7) | 74.1 [67.8-83.8] (60) | 0.721 |
| Remaining activities | 76.7 [71.2-85.0] (21) * | 83.8 [77.0-96.2] (110) * | *0.019* |

**Supplementary Table 2.** Heart rate variability (HRV) during daily life activities. Median [interquartile range] (number of participants).

|  | **Healthy controls** | **Long COVID** | *P* value |
| --- | --- | --- | --- |
|  | HRV (rMSSD) | HRV (rMSSD) |  |
| Sleep | 45.2 [28.6-61.0] (21) * | 32.6 [24.3-46.9] (119) * | *0.027* |
| House chores | 23.6 [17.6-34.6] (18) | 20.9 [15.1-26.6] (108) | 0.140 |
| Eating | 29.4 [19.3-41.7] (20) * | 22.5 [17.2-29.8] (110) * | *0.005* |
| Working | 31.2 [20.0-37.4] (20) | 25.6 [20.3-33.2] (83) | 0.239 |
| Traveling | 29.6 [20.6-37.5] (14)* | 22.4 [16.0-28.4] (99)* | *0.007* |
| Relaxing | 35.5 [16.9-47.4] (16) | 27.8 [19.2-39.0] (108) | 0.240 |
| Watching TV | 26.7 [19.1-43.5] (14) | 25.1 [20.0-35.3] (106) | 0.365 |
| Walking | 17.0 [11.5-23.7] (13) | 15.6 [10.9-20.5] (102) | 0.593 |
| Exercising | 12.4 [8.9-17.1] (21) | 13.4 [10.5-18.0] (113) | 0.253 |
| Getting ready | 24.0 [17.1-34.4] (7) | 23.8 [16.3-29.5] (57) | 0.639 |
| Relaxing exercise | 37.8 (1) | 26.1 [20.3-40.3] (54) | 0.179 |
| Reading | 20.3 [14.5-45.4] (7) | 26.2 [19.6-33.7] (73) | 0.647 |
| Remaining activities | 32.2 [19.7-43.5] (21) * | 24.2 [18.8-32.8] (120) * | *0.016* |

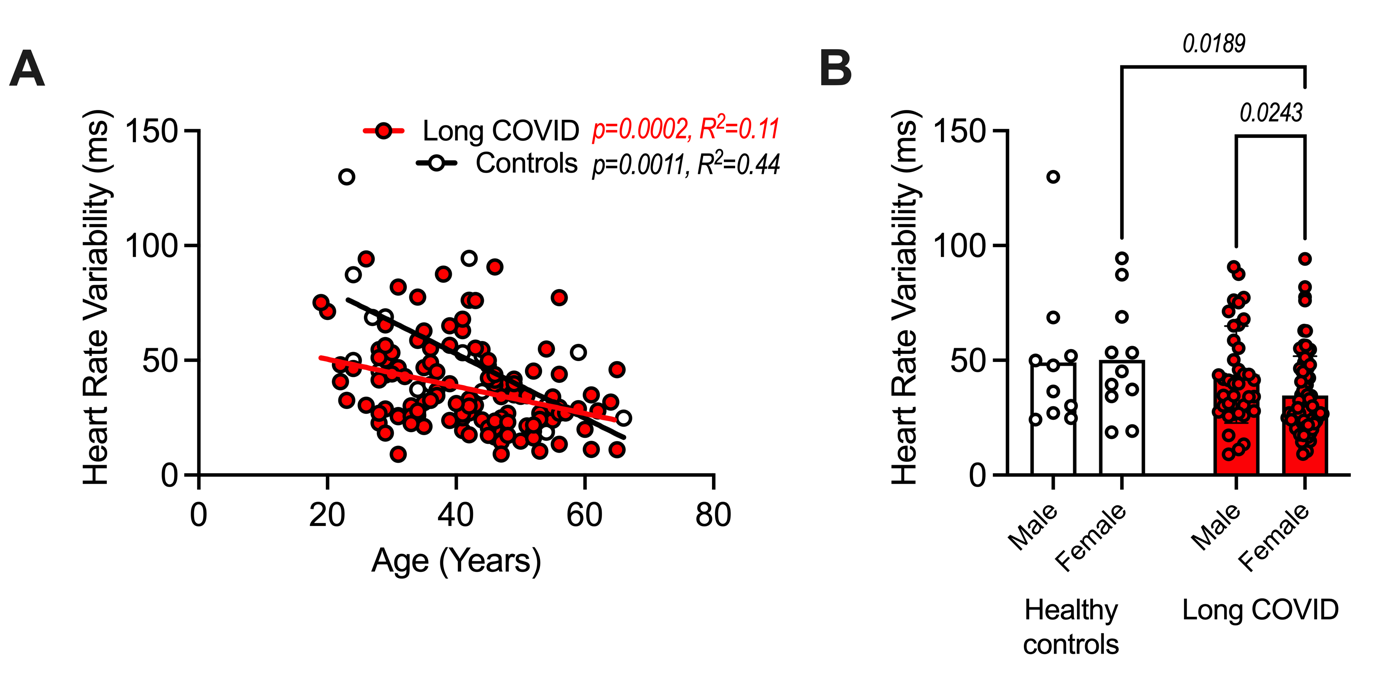

**Supplementary Figure 1.** (**A**) HRV was negatively associated to age in both patients with long Covid (p=0.0002, R^2^=0.11) and healthy controls (p=0.0011, R^2^=0.44). (**B**) Within the group of patients with long COVID, HRV was significantly lower in females (p=0.0243). Therefore, HRV statistical tests were corrected for age and sex.

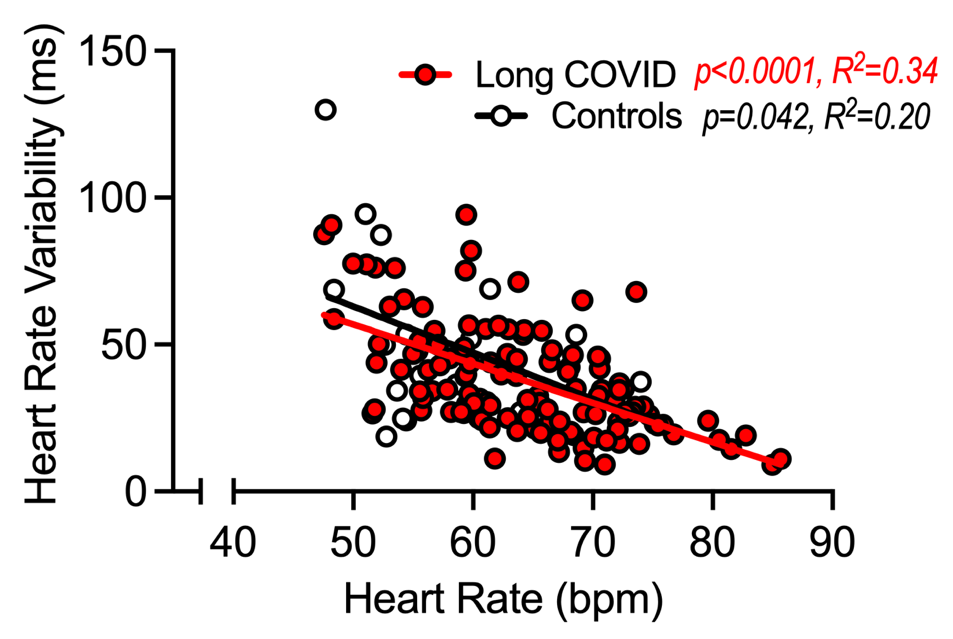

**Supplementary Figure 2.** Heart rate variability (HRV) significantly correlated to heart rate (HR) in both long COVID (p<0.0001, R^2^=0.34) and healthy controls (p=0.042, R^2^=0.20).
